# Correlation of Breast cancer Prognostic and Predictive indicators in Saudi patients: A 20 years retrospective observational study

**DOI:** 10.1101/2020.09.22.20173955

**Authors:** Haitham Kussaibi

## Abstract

**Introduction:** Breast cancer is the most common cancer in women in Saudi Arabia and the world (WHO 2020). Several studies have been published, worldwide, about the prognostic indicators of breast cancer, many of them showed a characteristic distribution according to certain geographical areas

**Methods:** Over 20 years (1998-2018), the results of 498 patients diagnosed with breast cancer, have been collected from the archive of the pathology department at King Fahd University Hospital (KFHU).

**Results:** This study included 498 patients diagnosed with breast cancer at King Fahd University Hospital over 20 years period (1998-2018), 58.4% (n=291) were Saudis. Data analysis showed a wide age distribution of breast cancers among eastern Saudi patients; however, most cases were seen in the 3rd and 4th decades. Luminal B was the most common subtype followed by triple-negative and luminal-A. Statistical analysis revealed a significant negative relationship between Saudi patients’ age at diagnosis and Her2 expression (P= .030), interestingly, this association was not significant in none-Saudi patients (P= .528).

**Discussion/Conclusion:** Our data revealed that breast cancer in Eastern Province had similar prognostics to international findings, however, Her2 profile and molecular subtype among Eastern Saudi women showed a minor deviation from worldwide published data.

## Introduction

Breast cancer is the most common cancer in women in Saudi Arabia constituting 18% of all cancers in Saudi women 1999 [1]. Several studies have been published, worldwide, about the prognostic indicators of breast cancer 2019-2020 [2-4], many of them showed a characteristic distribution according to certain geographical areas [5]. Breast cancer in Saudi Arabia has distinctive features than those in western countries 1998 [6]. The median age for Stage III breast cancers was 46 ± 11 which is distinct from the 60-65 years median age in Western countries 1999 [1]. The Eastern region of Saudi Arabia has the highest overall age-standardized incidence rate (26.6 per 100,000 women) 2013 [7]. A study in the western region included 740 cases, concluded that luminal A molecular subtype was the most common, while HER2-positive was the least, furthermore, HER2-positive and triple-negative tumors affected women below 50 years old and were associated with higher histological grade and larger tumor size 2020 [8], almost the same findings were reported in a study from Riyadh region on 359 breast cancer patients between 2010 and 2014 [9]. A 5 years study in the Eastern province between 1982-1987 revealed that breast cancer represented 4.1% of all malignancies diagnosed during this period 1991 [10]. An 8 years study (2006-2013) in Al Madinah region about histopathological features of breast cancer, showed that most cases were in an advanced stage at a young age (median 46) 2014 [11]. Another study in 2018 on 224 patients from Riyadh region, revealed a younger age distribution of breast cancer than those in Western nations, however, it was not associated with worse outcomes, overall survival for 10 years was 87% in all age groups [12]. In the Eastern province, there is a screening program in which annual screening is offered to all women aged 40 years and above. A study between 2009 and 2014 reported a total of 8061 women were screened representing 15.0%, cancer was detected in 47 (0.58%); of which 70% had no or less than 2 cm lesion [13]. A study between 1990 and 2014 revealed that breast cancer represented 27% of all female malignancies and it occurred at a younger age (median 48y) than those in western countries, there was a steady increase in cancer cases between 1990-2010 [14].

We aim here to reveal the profile of prognostic and predictive indicators of breast cancer from Saudi patients and none-Saudis living in the area and compare it with the published data worldwide and nearby region.

## Materials and Methods

### Data collection

Over 20 years (1998-2018), the results of 498 patients diagnosed with breast cancer, have been collected from the archive of pathology department at King Fahd University Hospital (KFHU).

### Inclusion criteria

All patients having breast cancer excisional specimen (lumpectomy or mastectomy).

### Data collected

Patients’ age at diagnosis, nationality, tumor type, grade, size, lymph node metastasis, ER, PR, Her2, Ki67, and molecular type. Results classified as the following:

Age at diagnosis: classified by decades. Patients’ nationality was either Saudi or non-Saudi. Tumor types included Ductal (NOS), Lobular, DCIS, or other. Concerning the tumor grading, we have followed the modified SBR system (grade I, II, or III), while for tumor size categorization, we have followed the TNM system (T1: ≤2cm, T2: 2-5cm, T3: >5cm). Lymph node status was either free or metastatic. ER & PR results were classified into three categories: 0, 1+ (1-10%), 2+ (11-50%), 3+ (> 50%). Her2 results grouped into positive (when it is 3+ by IHC or FISH) or negative (when it is 0 or 1+). The guidelines of the College of American Pathologists (CAP) has been adapted for ER, PR, and Her2. Ki67 results reported as neg, low (1-10%), intermediate (10-20%), or high proliferative index (> 20%). Molecular typing was according to 4 categories (luminal A, luminal B, triple-negative, or Her2).

### Data analysis

IBM SPSS v26 software has been utilized for statistical analysis. A significant correlation is considered at P-value ≤ *.050*.

## Results

### The demographic background of patients in the study

This study included 498 patients diagnosed with breast cancer at King Fahd University Hospital over 20 years period (1998-2017), 58.4% (n=291) were Saudis, while 42.6% (n=207) were non-Saudis. The none-Saudi patients included 36 Filipinos, 35 Egyptians, 21 Yemenis, 18 Syrians, 16 Sudani, 14 Jordanians, 12 Pakistanis, 11 Indonesians, 9 Indians, 6 Palestinians, 6 Lebanese, 4 Bahraini and 19 patients from other countries. The patients, either Saudi or none, come from different cities in the Eastern Province, mainly from Khobar and Dammam followed by Qatif and Hufuf then Dhahran, Hafr Albatin, and Jubail (as shown in Fig. 1.).

**Figure 1.**
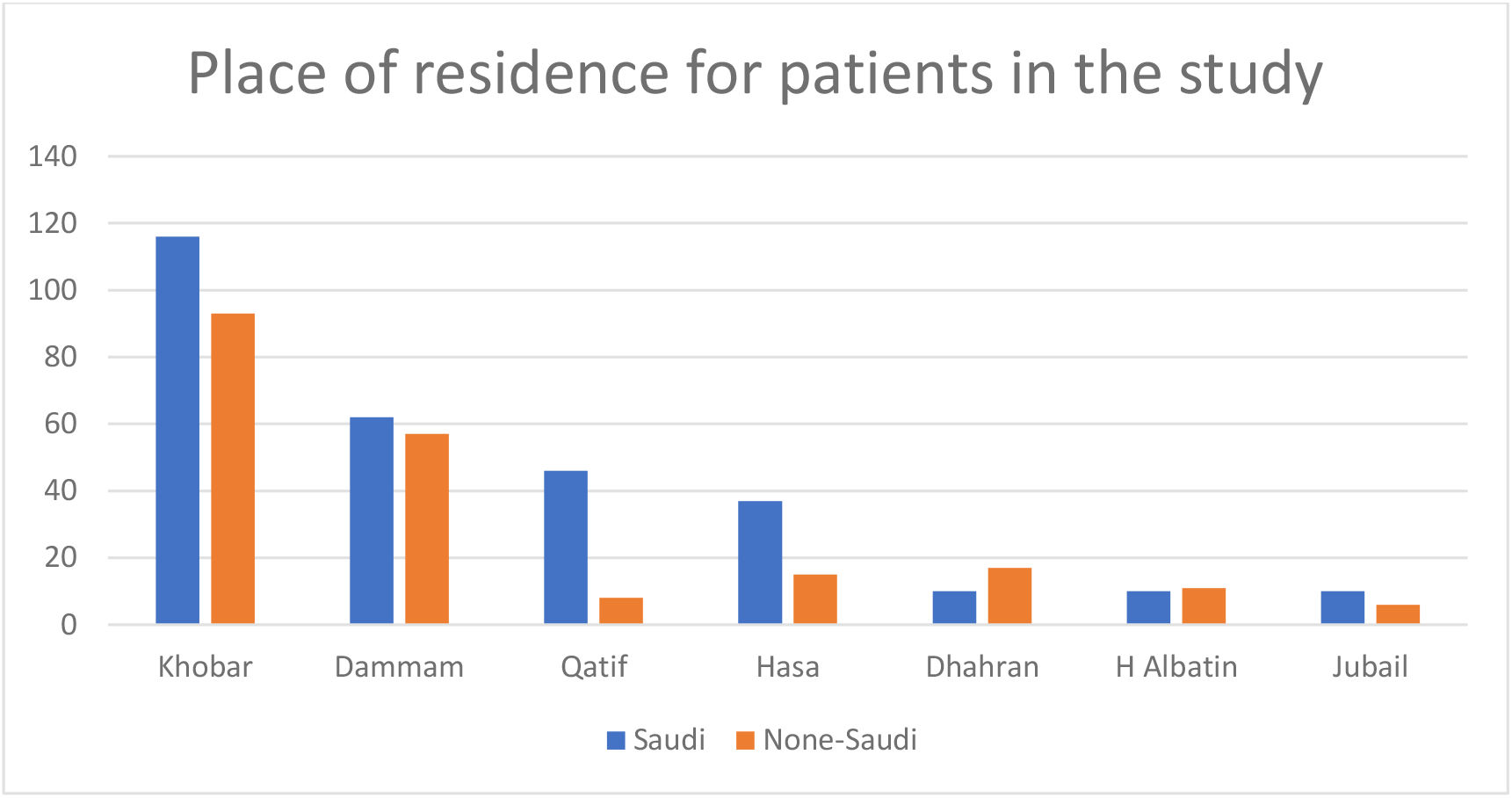
Place of residence for patients in the study

Over the 20 years of the study, there was a steady increase in the number of cancers diagnosed per year. Data analysis revealed an average of 11 new cases/year for the period (1998-2007) versus 39 new cases/year for the period (2008-2017).

Data analysis showed a wide age distribution of patients in the study, however, most breast cancer cases (76%) were seen in the 3rd, 4^th^’ and 5^th^ decades (as shown in Table 2). The median age at diagnosis was similar between Saudi and non-Saudi breast cancer patients (51y ± 12).

**Figure 2.**
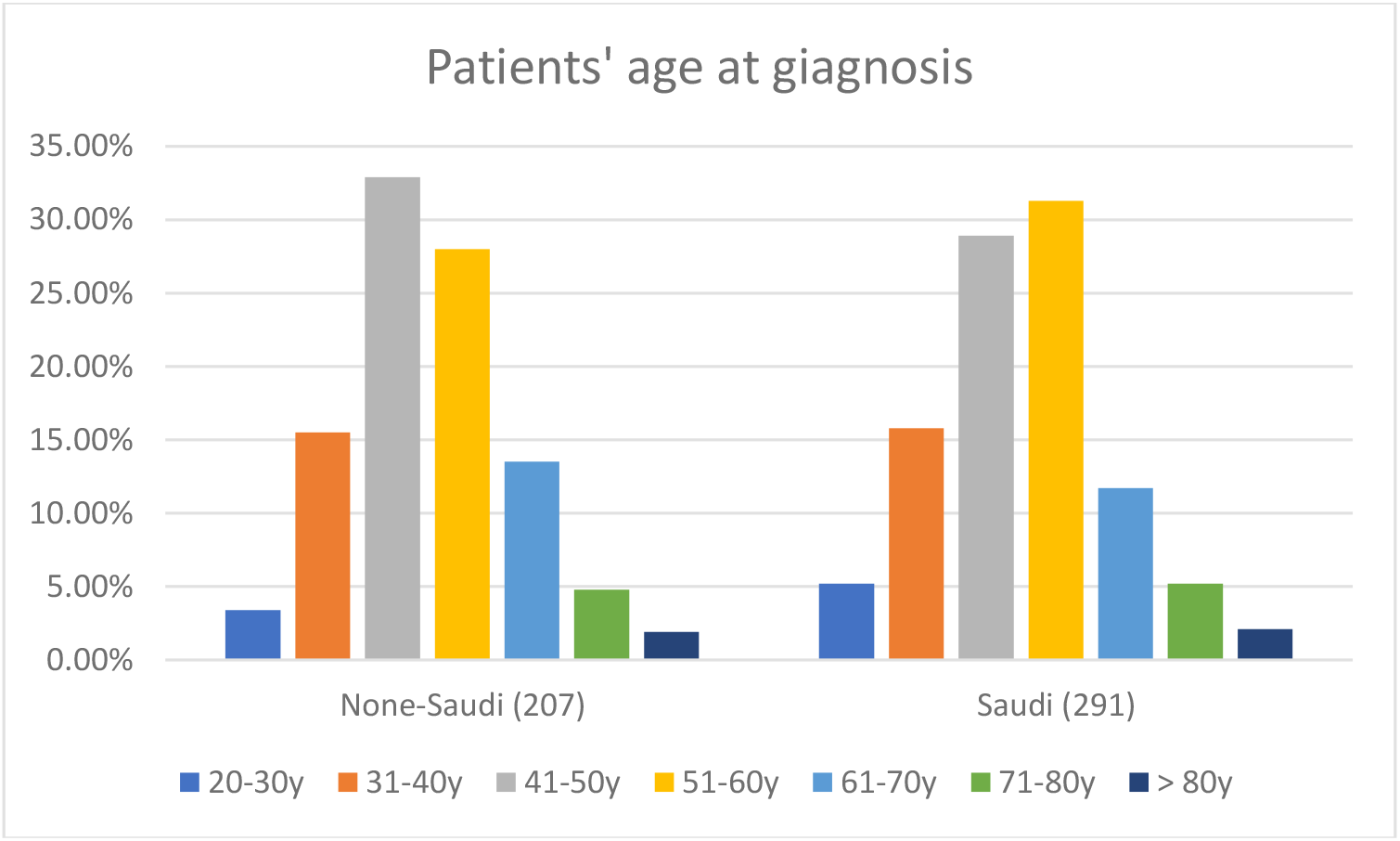
Patients’ age at diagnosis of breast cancer: comparison between Saudi and non-Saudi

**Table 1.** Frequency of breast cancer type, grade, and size among Saudi and non-Saudi patients:

|  | Cancer Type |  |  |  | Cancer Grade |  |  | Cancer size (cm) |  |  |
| --- | --- | --- | --- | --- | --- | --- | --- | --- | --- | --- |
|  | Ductal | Lobular | DCIS | Other | I/III | II/III | III/III | T1 (≤2) | T2 (>2 - ≤5) | T3 (>5cm) |
| Saudis | 258 (88.7%) | 20 (6.9%) | 5 (1.7%) | 8 (2.7%) | 24 (8.2%) | 159 (54.6%) | 108 (37.1%) | 34 (37.4%) | 43 (47.3%) | 14 (15.4%) |
| None-Saudis | 183 (88.4%) | 9 (4.3%) | 10 (4.8%) | 5 (2.4%) | 16 (7.7%) | 119 (57.5%) | 72 (34.8%) | 16 (23.2%) | 40 (58%) | 13 (18.8%) |

**Table 2.** Prognostic and predictive markers and molecular type for cancer patients in the study:

|  | ER status |  |  |  | PR status |  |  |  | Her2 status |  | Ki67 |  |  | Molecular type |  |  |  |
| --- | --- | --- | --- | --- | --- | --- | --- | --- | --- | --- | --- | --- | --- | --- | --- | --- | --- |
|  | -ve | +ve | +ve | +ve | -ve | +ve | +ve | +ve | -ve | +ve | Low | Intermediate | High | Luminal | Luminal | Triple- | Her2 |
|  |  | 1-10% | 11-50% | >50% |  | 1-10% | 11-50 | >50% |  | (3+) | 1-10% | 11-20% | 21-100% | A | B | negative |  |
| **Saudi** | 121 | 11 | 50 | 103 | 155 | 52 | 24 | 53 | 205 | 70 | 61 | 22 | 100 | 70 | 109 | 73 | 39 |
|  | (42.5%) | (3.9%) | (17.5%) | (36.1%) | (54.6%) | (18.3%) | (8.5%) | (18.7%) | (74.5%) | (25.5%) | (33.3%) | (12%) | (54.6%) | (24.1%) | (37.5%) | (25.1%) | (13.4%) |
| **Non-Saudi** | 65 | 4 | 28 | 48 | 83 | 27(18.6%) | 5 | 30 | 108 | 33 | 30 | 7 (8.1%) | 49 | 36 | 48 | 47 | 13 |
|  | (44.8%) | (2.8%) | (19.3%) | (33.1%) | (57.2%) |  | (3.4%) | (20.7%) | (76.6%) | (23.4%) | (34.9%) |  | (57%) | (25%) | (33.3%) | (32.6%) | (9%) |

### Histopathological features of breast cancers in the study

In Saudi patients, invasive ductal carcinoma (NOS) was the most common type (88.7%), followed by invasive lobular carcinoma (6.9%), DCIS (1.7%), and other types (2.7%). Non-Saudis showed a comparable frequency of the different types; however, DCIS was slightly more frequent (4.8%) (shown in **Table 1**.).

Most tumors were of grade II/III (54.6%), followed by Grade III/III (37.1%). Grade I/III was the least seen (8.2%). The results were again almost similar between Saudis and non-Saudis.

One third (37.4%) of resected tumors in Saudi patients, were in stage T1, while 54.6% were in stage T2, however, only 15.4% were in advanced stage (T3). Tumors’ stage was slightly different in non-Saudi patients included 23.2% in stage T1, 58% in stage T2, and 18.8% in stage T3.

For patients having axillary dissection, lymph node metastasis was seen in 56.6% and 67% of Saudi and non-Saudi patients, respectively.

### Prognostic and predictive markers along with molecular type for breast cancer cases in the study

The negativity rate in Saudi patients for ER, PR, and Her2 was 42.5%, 54.6%, and 74.5%, respectively. Comparable results were found in non-Saudis.

In 2 thirds of cancers, the Ki67 proliferative index was more than 10%.

Molecular type luminal B was the most common among Saudi patients representing 37.5%, followed by triple-negative and luminal A, which represent around 25% each. The lease common type was Her2 type (13.4%). Similar findings were noticed for non-Saudi patients except for triple-negative type which was slightly more frequent (32.6%) than that in Saudi patients.

### Statistical analysis

Correlation analysis by IBM SPSS v26 using Spearman and Pearson revealed a negative significant relationship between Saudi patients’ age at diagnosis and Her2 expression (P= .030) which means, Her2 amplification/high expression was more frequently seen in relatively younger patients. Interestingly, this association was not significant in none-Saudi patients (P= .528). Furthermore, it has been observed that breast cancer, over the years, is diagnosed more and more in younger ages, this observation was also in Saudi patients rather than none-Saudis which has also been confirmed by correlation analysis (P= .002 in Saudis, P= .267 in none-Saudis). In addition to that, over the years between 1998 and 2017, it has been noticed that breast cancers are more and more of luminal types (P= .000) with lower grades (P= .012) and lower stage (less metastasis to lymph nodes) (P= .009). This last observation, however, was comparable between Saudis and none-Saudis.

## Discussion/Conclusion

Our data revealed that breast cancer in Eastern Province had similar prognostics to international findings, however, Her2 profile and molecular type were a little bit different and showed characteristic distribution among Saudi women in this region.

The distribution of patients among the different cities is logical because of the hospital location in Al-Khobar city so most of the patients live in that city in addition to its very close neighbor cities Dammam and Dhahran.

Over the 20 years of our study, there was a steady increase in cancer incidence rate per year. A similar observation has been reported in a study by Saggu et al, based on 15 years (1990-2014) data obtained from the Saudi cancer registry. This is also reported, worldwide, in most countries mainly in developing countries, as shown in a recent study by [15] (based on data collected between 1990 - 2017), revealed that female breast cancers increased from 870.2 thousand to 1937.6 thousand, with age-standardized incidence rate increased from 39.2/100,000 to 45.9/100,000, mainly found in developing countries, however, many developed countries (like USA and UK) showed decrease in incidence rate.

Most cancer cases (76%), in the study, were seen in the 3rd - 5th decades of the patient’s life, the median age at diagnosis was (51y ± 12) and there was no difference between Saudi or non-Saudi patients. This average age is a little bit higher than several studies in Saudi Arabia, for example, a study by Alnegheimish NA et al done on 359 patients with BC from Riyadh region revealed a mean age of 49.8 years [9]. Another study by Abdulkader Albasri et al on 398 patients with BC from the Almadinah region, revealed a mean age of 46.9 years [11]. Other middle-eastern countries such as

Lebanon, showed almost similar mean age, as mentioned in a Lebanese study by Nagi S El Saghir et al who reported a mean age of 49.8 years +/- 13.9 years among 2673 female with breast cancers [16]. However, this is far younger than the mean age of breast cancer patients reported in developed countries such as the USA and the UK which is around 60 -65 years [6].

Invasive ductal carcinoma (NOS) was the most common type (88.7%), followed by invasive lobular carcinoma (6.9%), DCIS (1.7%), and other types (2.7%). These findings are comparable to several studies in Saudi Arabia and worldwide [3, 11]. Most tumors were of grade II/III (54.6%), followed by Grade III/III (37.1%). Grade I/III was the least seen (8.2%). This is also reported similarly in several studies in Saudi Arabia and worldwide.

One third (37.4%) of resected tumors were in stage T1, while 54.6% were in stage T2, however, only 15.4% were in advanced stage (T3). Tumors in stage T1 were slightly different in non-Saudi patients included 23.2% vs 37.4%., this could be explained by more accessibility of Saudis to cancer screening which increases the detection of cancer in an earlier stage. Furthermore, T2 (2-5cm) tumors were the most frequent in many studies in Saudi Arabia and other developing countries, while, T1 tumors are the most frequent ones in developed countries, again, due to the earlier detection of cancers, according to many studies around the globe.

For patients having axillary dissection, lymph node metastasis was seen in 56.6% and 67% of Saudi and non-Saudi patients, respectively. In general, cancers in none-Saudis were more advanced in stage, not only in tumor size but also in cancer metastasis, either to lymph nodes or to distant organs. This is most probably, as mentioned above, due to the late detection of cancers for none-Saudi, as they have less access to health services than native citizens.

The negativity rate in Saudi patients for ER, PR, and Her2 was 42.5%, 54.6%, and 74.5%, respectively. According to the College of American Pathologists (CAP) guidelines, the overall proportion of ER-negative breast cancers (invasive and DCIS) in general, should not exceed 30% (approximately 20% for postmenopausal vs. 35% for premenopausal women and up to 10% in Grade I tumors), while, the overall proportion of HER2 positive breast cancers is 10-25% [17], [18]. In our series, the proportion ER-negative breast cancers is much higher (42.5%), this is logic when we know that 92% of cancers in our series are of Grade II or Grade III which are less frequently expressing ER or PR receptors, however, the positivity rate of Her2 was 25.5% in our study which is within the published benchmarks.

In 2 thirds of cancers, the Ki67 proliferative index was more than 10% which is well correlated with the high frequency of Grade II (54.6%) and Grade III (37.1%) cancers in our series. A study from the Al-Madinah region reported (Ki67 > 25%) in 73.9% of the 115 included patients [19]. However, there is variability among pathologists about the cut-off value of High Ki67 [20].

Molecular type luminal-B was the most common among Saudi patients representing 37.5%, followed by triple-negative and luminal-A, which represent around 25% each. The least common type was Her2 type (13.4%). This is different from a Saudi study in the Riyadh region on 359 patients which found that Luminal-A was the most common type (58.5%), triple-negative (14.8%), luminal-B (14.5%), and Her2+ (12.3%) [9]. Similarly, a study from Oman on 542 breast cancer cases revealed that Luminal-A was the most common subtype followed by triple-negative subtype. [5]. The higher frequency of luminal-B in our series could be due-to considering high Ki67, in addition to the positive receptor (ER or PR), as a qualifier to classify cancers as luminal-B even with negative Her2.

It has been revealed a negative significant relationship between Saudi patients’ age at diagnosis and Her2 expression (P= .030) which means, Her2 amplification/high expression was more frequently seen in younger patients. One third of Her2-positive Saudi breast cancer patients were in their 3^rd^ or 4^th^ decade of life at diagnosis, while only 16% of patients with Her2-negative breast cancer were at these age period (3^rd^ - 4^th^ decade). In other word, 41% of breast cancer in young age (3^rd^ - 4^th^ decade) were Her2-positive. This is quitely like a Japanese study on 25898 BC patients which revealed that young patients (< 35 years) were associated with an advanced TNM stage and aggressive characteristics including Her2-positivity, compared to older patients [21]. Interestingly, this association was not significant in none-Saudi patients (P= .528). However, a study in Jeddah on 145 patients revealed no significant correlations between HER-2/neu and age or race [22].

Furthermore, it has been observed that breast cancer, over the years, is diagnosed more and more in younger ages, this observation was also in Saudi patients rather than none-Saudis which has also been confirmed by correlation analysis (P= .002 in Saudis, P= .267 in none-Saudis). In addition to that, over the years between 1998 and 2017, it has been noticed that breast cancers are more and more of luminal types (P= .000) with lower grade (P= .012) and lower stage (less metastasis to lymph nodes) (P= .009). This could be explained by the improvement of early detection of cancer over years.

Our data revealed that breast cancer in Eastern Province had similar prognostics to international findings, however, Her2 profile and molecular subtype among Eastern Saudi women showed a minor deviation from worldwide published data.

## Statements

## Data Availability

Data are not available for public use.

## Acknowledgement

The author is grateful to Dr Areej Al Nemer, Pathology associate professor and consultant at King Fahd Hospital, Imam Abdulrahman Bin Faisal University, for her valuable support and feedback.

## Statement of Ethics

Ethical approval is granted by the SCRELC (Standing Committee on Research Ethics on Living Creatures), Institutional review board at Imam Abdulrahman Bin Faisal University, IRB-2020-1-262.

Informed patient consent was waived due to the use of archival data of anonymous nature that does not disclose patients’ identity.

## Author Contributions

This work has been conducted by a single author.

## References

1. Ezzat AA, Ibrahim EM, Raja MA, Al-Sobhi S, Rostom A, Stuart RK. Locally advanced breast cancer in Saudi Arabia: high frequency of stage III in a young population. Med Oncol. 1999;16(2):95–103.

2. Babiker S, Nasir O, Alotaibi SH, Marzogi A, Bogari M, Alghamdi T. Prospective breast cancer risk factors prediction in Saudi women. Saudi J Biol Sci. 2020;27(6):1624–31.

3. Asiri S, Asiri A, Ulahannan S, Alanazi M, Humran A, Hummadi A. Incidence Rates of Breast Cancer by Age and Tumor Characteristics Among Saudi Women: Recent Trends. Cureus. 2020;12(1):e6664.

4. Alsolami FJ, Azzeh FS, Ghafouri KJ, Ghaith MM, Almaimani RA, Almasmoum HA, et al. Determinants of breast cancer in Saudi women from Makkah region: a case-control study (breast cancer risk factors among Saudi women). BMC Public Health. 2019;19(1):1554.

5. Mehdi I, Monem AA, Al Bahrani B, Ramadhan FA. Breast cancer molecular subtypes in oman: correlation with age, histology, and stage distribution - analysis of 542 cases. Gulf J Oncolog. 2014;1(15):38–48.

6. Ibrahim EM, al-Mulhim FA, al-Amri A, al-Muhanna FA, Ezzat AA, Stuart RK, et al. Breast cancer in the eastern province of Saudi Arabia. Med Oncol. 1998;15(4):241–7.

7. Alghamdi IG, Hussain, II, Alghamdi MS, El-Sheemy MA. The incidence rate of female breast cancer in Saudi Arabia: an observational descriptive epidemiological analysis of data from Saudi Cancer Registry 2001-2008. Breast Cancer (Dove Med Press). 2013;5:103-9.

8. Al-Thoubaity FK. Molecular classification of breast cancer: A retrospective cohort study. Ann Med Surg (Lond). 2020;49:44-8.

9. Alnegheimish NA, Alshatwi RA, Alhefdhi RM, Arafah MM, AlRikabi AC, Husain S. Molecular subtypes of breast carcinoma in Saudi Arabia. A retrospective study. Saudi Med J. 2016;37(5):506–12.

10. al-Idrissi HY. Pattern of breast cancer in Saudi females in eastern province of Saudi Arabia. Indian J Med Sci. 1991;45(4):85–7.

11. Albasri A, Hussainy AS, Sundkji I, Alhujaily A. Histopathological features of breast cancer in Al-Madinah region of Saudi Arabia. Saudi Med J. 2014;35(12):1489–93.

12. Alabdulkarim B, Hassanain M, Bokhari A, AlSaif A, Alkarji H. Age distribution and outcomes in patients undergoing breast cancer resection in Saudi Arabia. A single-institute study. Saudi Med J. 2018;39(5):464–9.

13. Al Mulhim FA, Syed A, Bagatadah WA, Al Muhanna AF. Breast cancer screening programme: experience from Eastern province, Saudi Arabia. East Mediterr Health J. 2015;21(2):111–9.

14. Saggu S, Rehman H, Abbas ZK, Ansari AA. Recent incidence and descriptive epidemiological survey of breast cancer in Saudi Arabia. Saudi Med J. 2015;36(10):1176–80.

15. Chen Z, Xu L, Shi W, Zeng F, Zhuo R, Hao X, et al. Trends of female and male breast cancer incidence at the global, regional, and national levels, 1990-2017. Breast Cancer Res Treat. 2020;180(2):481–90.

16. El Saghir NS, Shamseddine AI, Geara F, Bikhazi K, Rahal B, Salem ZM, et al. Age distribution of breast cancer in Lebanon: increased percentages and age adjusted incidence rates of younger-aged groups at presentation. J Med Liban. 2002;50(1-2):3-9.

17. Allison KH, Hammond MEH, Dowsett M, McKernin SE, Carey LA, Fitzgibbons PL, et al. Estrogen and Progesterone Receptor Testing in Breast Cancer: ASCO/CAP Guideline Update. J Clin Oncol. 2020;38(12):1346–66.

18. Wolff AC, Hammond MEH, Allison KH, Harvey BE, Mangu PB, Bartlett JMS, et al. Human Epidermal Growth Factor Receptor 2 Testing in Breast Cancer: American Society of Clinical Oncology/College of American Pathologists Clinical Practice Guideline Focused Update. J Clin Oncol. 2018;36(20):2105–22.

19. Elkablawy MA, Albasri AM, Mohammed RA, Hussainy AS, Nouh MM, Alhujaily AS. Ki67 expression in breast cancer. Correlation with prognostic markers and clinicopathological parameters in Saudi patients. Saudi Med J. 2016;37(2):137–41.

20. Al Nemer A. The Performance of Ki-67 Labeling Index in Different Specimen Categories of Invasive Ductal Carcinoma of the Breast Using 2 Scoring Methods. Appl Immunohistochem Mol Morphol. 2017;25(2):86–90.

21. Kataoka A, Iwamoto T, Tokunaga E, Tomotaki A, Kumamaru H, Miyata H, et al. Young adult breast cancer patients have a poor prognosis independent of prognostic clinicopathological factors: a study from the Japanese Breast Cancer Registry. Breast Cancer Res Treat. 2016;160(1):163–72.

22. Al-Ahwal MS. HER-2 positivity and correlations with other histopathologic features in breast cancer patients--hospital based study. J Pak Med Assoc. 2006;56(2):65–8.

